# Rapid detection of single nucleotide polymorphisms using the MinION nanopore sequencer: a feasibility study for perioperative precision medicine

**DOI:** 10.1101/2022.01.13.22269267

**Authors:** Yoshiteru Tabata, Yoshiyuki Matsuo, Yosuke Fujii, Atsufumi Ohta, Kiichi Hirota

## Abstract

**Introduction:** Precision medicine is a phrase used to describe personalized medical care tailored to specific patients based on their clinical presentation and genetic makeup. However, despite the fact that several single nucleotide polymorphisms (SNPs) have been reported to be associated with increased susceptibility to particular anesthetic agents and the occurrence of perioperative complications, genomic profiling and thus precision medicine has not been widely applied in perioperative management.

**Methods:** We validated six SNP loci known to affect perioperative outcomes in Japanese patients using genomic DNA from saliva specimens and nanopore sequencing of each SNP loci to facilitate allele frequency calculations and then compared the nanopore results to those produced using the conventional dideoxy sequencing method.

**Results:** Nanopore sequencing reads clustered into the expected genotypes in both homozygous and heterozygous cases. In addition, the nanopore sequencing results were consistent with those obtained using conventional dideoxy sequencing and the workflow provided reliable allele frequency estimation, with a total analysis time of less than 4 h.

**Conclusion:** Thus, our results suggest that nanopore sequencing may be a promising and versatile tool for SNP genotyping, allowing for rapid and feasible risk prediction of perioperative outcomes.

## Introduction

The most critical role of any anesthesiologist is to ensure the safety of their patient during the perioperative period and provide the skills and expertise to ensure a smooth surgical experience. The patients’ medical conditions can be affected by a complex combination of factors, including the degree of invasiveness of the surgery, the effects of the anesthesia, and the reserve capacity of the patient. A preoperative medical assessment is used to ensure the safety and comfort of patients undergoing surgery. However, even with preoperative testing, it is difficult to predict individual susceptibility to particular anesthetic drugs and the risk of potential postoperative complications such as postoperative nausea and vomiting [1, 2].

Precision medicine is an emerging approach designed to facilitate improved prevention and treatment of human diseases [3]. This approach uses personalized medical care tailored to a specific patient based on their clinical presentation and genetic background to improve clinical outcomes. In cancer treatment, precision medicine uses genetic information derived from the patient’s tumor cells to help diagnose and select appropriate targeted therapies [4, 5] and although the utility of precision medicine continues to be increasingly accepted across a wide range of medical fields, its application in perioperative management remains limited.

Previous studies have shown that there are a range of specific genetic variations associated with changes in susceptibility to specific anesthetic agents and perioperative outcomes [6]. Single nucleotide variations (SNV) are characterized by an alteration at a single position within a DNA sequence and when this SNV is present in at least 1% of the population, it is referred to as a single nucleotide polymorphism (SNP). For instance, SNP rs1799971, which is located in the gene encoding opioid receptor mu 1 (*OPRM1*), is a well understood genetic variation with significance for perioperative management. The rs1799971 A>G substitution results in an asparagine-to-aspartate conversion at position 40 of the protein, leading to increased opioid resistance and thus a need for an increase in therapeutic dose [7-9]. Along with recent advances in DNA sequencing technology, the development of an integrated SNP dataset would be beneficial for the prediction of potential perioperative risk and the management of anesthetic complications.

Given this we designed this study to evaluate a streamlined workflow for SNV/SNP genotyping using nanopore sequencing technology, with a view to enabling genetic based perioperative risk evaluations in preoperative patients. The MinION nanopore sequencer is a portable DNA/RNA sequencing platform that provides on-site genetic analysis with rapid and affordable deployment [10, 11]. We developed a simple bioinformatics pipeline to reliably detect several SNPs of interest within the nanopore sequencing data. We selected six SNP loci affecting perioperative outcomes for validation, and the utility of the workflow was evaluated using SNP allele frequency estimations and their comparison with more conventional sequencing technologies.

## Methods

### Samples and DNA extraction

This study was approved by the institutional review board at Kansai Medical University Hospital (No. 2020285, February 19, 2021) and saliva samples were collected from five willing participants. Approximately 1 mL of saliva was mixed with 4 mL of phosphate-buffered saline (PBS, Nacalai Tesque, Kyoto, Japan) and centrifuged at 1800 × g for 5 min before being resuspended in 0.3 mL of PBS and subjected to DNA extraction using the Maxwell RSC Blood DNA Kit (AS1400, Promega, Madison, WI, USA) on the Maxwell RSC automated nucleic acid purification platform (AS4500, Promega).

Briefly, the sample (0.3 mL) was mixed with lysis buffer (0.3 mL) and proteinase K solution (30 µL), incubated at 56 °C for 20 min, and transferred to a Maxwell RSC Cartridge for magnetic bead-based DNA extraction. DNA was eluted in 50 µL elution buffer and quantified using the QuantiFluor ONE dsDNA System (E4871, Promega).

### DNA amplification

A detailed protocol of the 2-step polymerase chain reaction (PCR) for nanopore amplicon sequencing is available at protocols.io (https://dx.doi.org/10.17504/protocols.io.bwr5pd86). Briefly, six SNP loci were used for amplicon sequencing (Table 1) and the locus-specific sequences of the PCR primers are listed in Table 2.

**Table 1.** List of SNPs evaluated in this study

| SNP | Gene | Full gene name | Clinical significance | Ref. |
| --- | --- | --- | --- | --- |
| rs1045642 | *ABCB1* | ATP binding cassette subfamily B member 1 | A>G: increased susceptibility to PONV<br>A>G: increased opioid dose requirement | 18<br>19 |
| rs1799971 | *OPRM1* | opioid receptor mu 1 | A>G: increased opioid dose requirement | 7-9 |
| rs2165870 | *CHRM3* | cholinergic receptor muscarinic 3 | A>G: decreased susceptibility to PONV | 20, 21 |
| rs4369876 | *SCN9A* | sodium voltage-gated channel alpha subunit 9 | C>A: decreased pain sensitivity | 22 |
| rs33985936 | *SCN11A* | sodium voltage-gated channel alpha subunit 11 | C>T: increased pain sensitivity | 23 |
| rs140124801 | *KCNG4* | potassium voltage-gated channel modifier subfamily G member 4 | C>T: decreased pain sensitivity | 24 |

PONV, postoperative nausea and vomiting

**Table 2.**
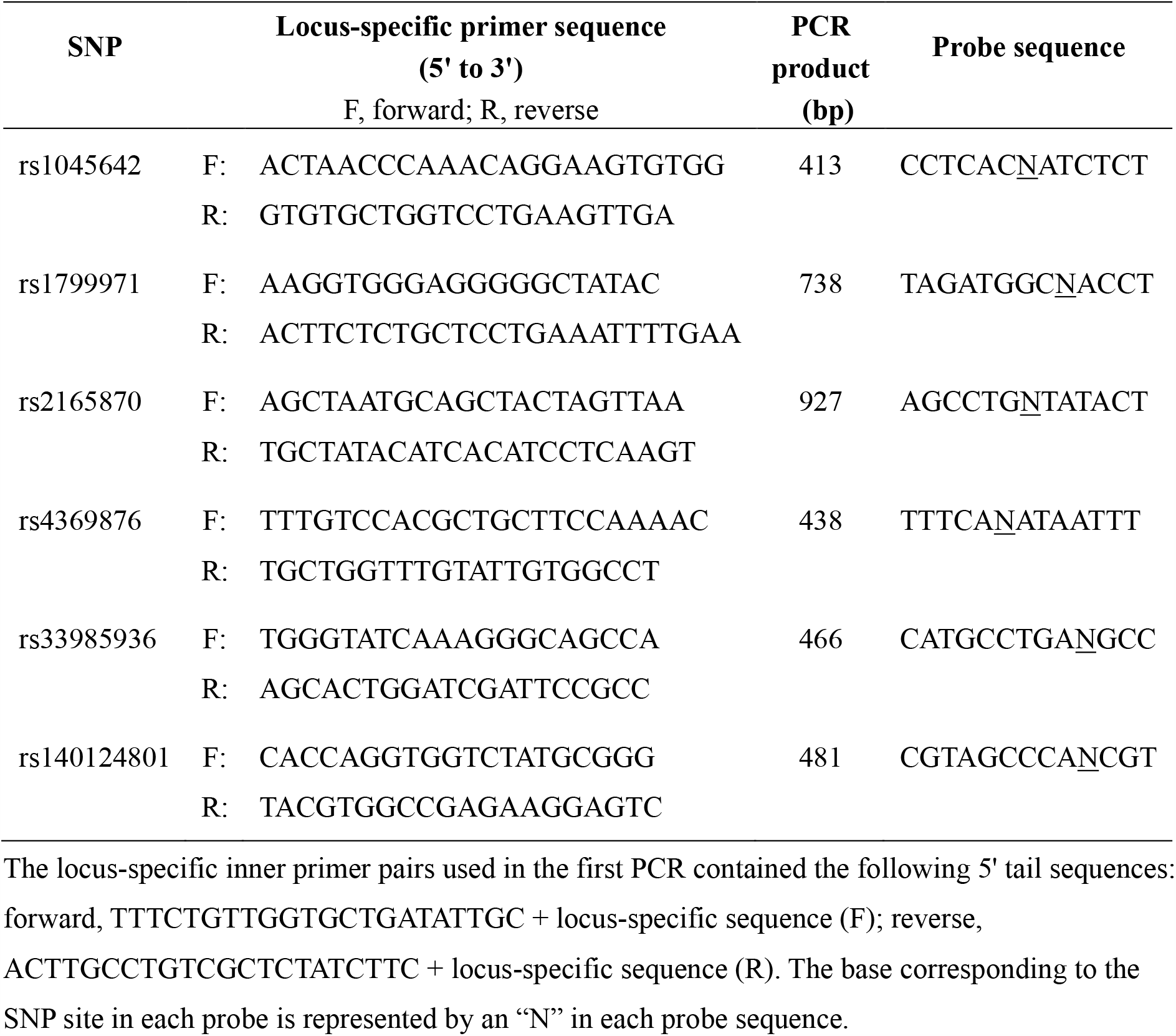
Primer and probe sequences used in the nanopore-mediated genotyping of the six target SNPs

A total of 20 ng of saliva DNA was used as a template to amplify the target genomic region and the locus-specific inner primers used in the first PCR each included the following 5’ tail sequences: forward 5’-TTTCTGTTGGTGCTGATATTGC - locus-specific sequence-3’; reverse 5’-ACTTGCCTGTCGCTCTATCTTC - locus-specific sequence-3’. PCR amplification was performed using Platinum II Hot-Start PCR Master Mix (14000012, Thermo Fisher Scientific, Waltham, MA, USA) with 0.2 µM of each inner primer in a total volume of 25 μL. Amplification conditions were as follows: initial denaturation at 94 °C for 2 min, 35 cycles of 94 °C for 15 s, 60 °C for 15 s, 68 °C for 30 s, followed by a final extension at 68 °C for 1 min. The resultant amplicons (1 µL) were subjected to a second PCR to introduce the indices (barcodes) and the 5’ tags required for adapter attachment. These reaction mixtures (25 µL) contained KAPA2G Robust HotStart ReadyMix (KK5701, KAPA Biosystems, Wilmington, MA, USA) and the barcoded outer primers (0.5 µL) supplied in the PCR Barcoding Kit (SQK-PBK004, Oxford Nanopore Technologies, Oxford, UK). Cycling conditions were as follows: initial denaturation at 95 °C for 3 min; 15 cycles of 95 °C for 15 s, 62 °C for 15 s, 72 °C for 30 s, followed by a final extension at 72 °C for 1 min. Amplified DNA was then purified using AMPure XP (A63880, Beckman Coulter, Brea, CA, USA) and quantified using a QuantiFluor ONE dsDNA System.

### Nanopore sequencing

The purified barcoded amplicons were then pooled, and 100 fmol was applied as template in the library preparation which was completed using the PCR Barcoding Kit. These libraries were loaded onto the R9.4.1 flow cell (FLO-MIN106, Oxford Nanopore Technologies) and sequenced on the MinION Mk1C with MinKNOW software version 21.05.21 (Oxford Nanopore Technologies). Base-calling was performed in real time via Guppy version 5.0.13 (Oxford Nanopore Technologies) using the following settings: fast basecalling model, trim_barcodes=on, require_barcodes_both_ends=off, detect_mid_strand_barcodes=on, min_score=60. Called reads (FASTQ format) were filtered to generate pass reads, with a minimum Phred quality score of 8.

### DNA sequencing using the dideoxy method

The first round PCR products described above were purified using an AMPure XP and sequenced on both strands using the following primers: forward TTTCTGTTGGTGCTGATATTGC and reverse ACTTGCCTGTCGCTCTATCTTC, which correspond with the 5’ tails described above. Sequencing was performed on a 3130xl Genetic Analyzer using a BigDye Terminator v3.1 Cycle Sequencing kit (Thermo Fisher Scientific).

### Bioinformatic analysis

The average Phred quality scores of the nanopore sequencing reads were analyzed using NanoPlot ver. 1.27.0 and the allele frequencies for each of the target SNP sites were determined using the following bioinformatics pipeline in SeqKit version 0.13.2. Step 1: Reads with a perfect match to the probe sequence specific to each SNP site (Table 2) were extracted and saved into individual files. Where this command identifies the probe sequence (SNP site is represented by “N”) in both the forward and reverse strands of the amplicon. Step 2: random sampling of one thousand reads per target. Step 3: Reads were sorted by SNP genotype, and allele frequencies were calculated based on the read count. The exact commands used to generate this data are provided in Additional file 1.

## Results

Variant data were obtained from the Integrated Genome Variation Database, TogoVar [12] (Additional file 2: Table S1) and then used to identify six SNP loci associated with specific perioperative outcomes that could be used to evaluate the feasibility of nanopore sequencing in the real-time analysis of genetic predisposition to adverse perioperative outcomes (Table 1). Genomic DNA was extracted from the saliva of five individuals and then subjected to PCR amplification of the target loci using specific primers (Table 2), before sequencing on the MinION platform. These amplicons were also sequenced using the dideoxy method to verify the genotypes (Additional file 3, Figs. S1-S6).

Nanopore sequencing was performed on up to 12 barcoded samples at a time, and 10-minutes of MinION sequencing yielded an average of over 30000 pass reads making this sequencing time sufficient for SNP genotyping using our bioinformatic pipeline. Nanopore sequencing reads with a perfect match to the probe sequence (Table 2) were sorted, and 1000 reads per sample were collected and then used to determine the allele frequencies of each SNP site (Additional file 4: Table S2). The estimation of the allele frequencies for the target SNP loci was completed in approximately 3.5 hours (Fig. 1). Allele frequencies as determined by nanopore sequencing fluctuated around the theoretical value of 50% for heterozygote variants. While in the homozygote samples, the reads were binned into only one of the given variants, with allele frequencies of approximately 100% (Fig. 2). A maximum deviation of 8.4% from the expected value was observed (Fig. 2c and Additional file 5, Table S5) and only a small fraction of false-positive variants were detected (< 6%; Fig. 2c and Additional file 5: Tables S3-S8).

**Fig. 1.**
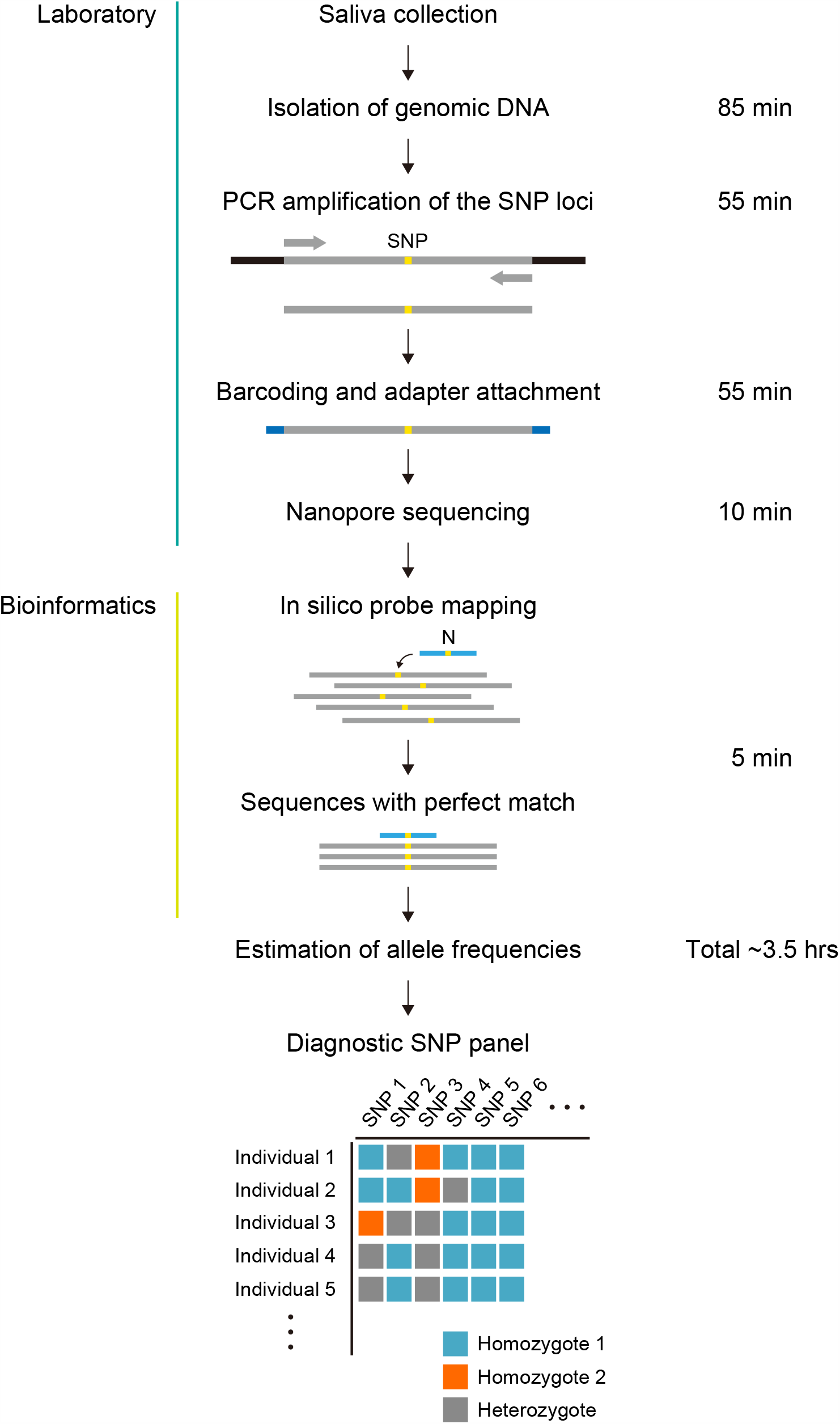
Schematic describing the laboratory and bioinformatics pipelines used to complete targeted SNP genotyping using the nanopore sequencing technology.

**Fig. 2.**
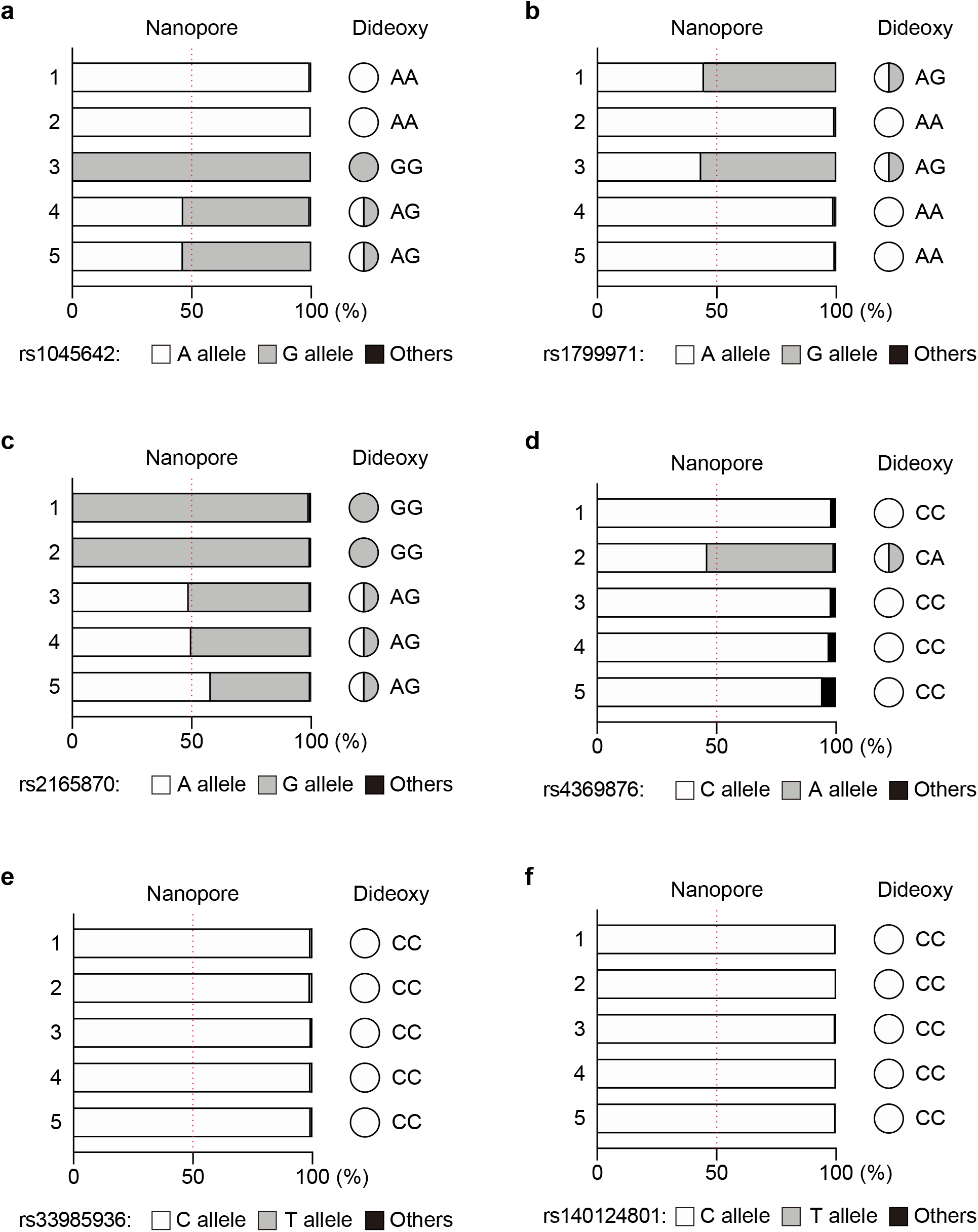
Genotyping of SNPs associated with perioperative outcomes and anesthetic complications. Six SNP loci were genotyped from five individuals: a rs1045642, b rs1799971, c rs2165870, d rs4369876, e rs33985936, f rs140124801. The graphs show the allele frequencies determined by nanopore amplicon sequencing. These amplicons were then sequenced using traditional dideoxy sequencing to confirm their genotypes.

Given that the mean Phred quality score of the nanopore reads was 12.2 (∼94% accuracy, Additional file 4: Table S2), we suggest that such spurious sequence variation is likely the result of minor sequencing errors. Taken together our results show that the nanopore sequencing reads are reasonably well clustered within the expected allele frequencies, demonstrating the reliability of this approach for SNP genotyping.

## Discussion

Recent technological advances in genomic research, including next-generation sequencing (NGS), have enabled rapid and precise analysis of large amounts of genomic data [13]. This has meant that genomic profiling is increasingly being used to identify subpopulations with different susceptibilities to particular drugs and medical treatments, facilitating better, more personalized treatment [14]. These integrated genomic datasets demonstrate significant potential for developing targeted medical strategies based on individual variability in specific patient characteristics. Here, we developed a novel nanopore-based sequencing strategy for rapid SNP genotyping and evaluated its feasibility for genomic profiling in the context of perioperative care, in an attempt to integrate precision medicine into this unique medical discipline.

Recent reports have described several genetic variations that may affect a patient’s susceptibility to particular anesthetic agents and the occurrence of perioperative complications [6]. These include SNP rs1799971 in *OPRM1* which has been extensively studied as an example of a genetic variant affecting perioperative outcomes. *OPRM1* encodes the µ-opioid receptor [15], which is the primary site of action for most perioperative opioids, including morphine and fentanyl. This SNP is located in the protein-coding region, and the 118A>G substitution causes an amino acid change (p.Asn40Asp), potentially affecting receptor expression and the downstream signaling pathway [16, 17]. It has also been suggested that patients with AG and GG genotypes require increased opioid doses for pain management [7-9]; however, the mechanisms underlying these potential associations remain unclear. In addition, while the overall frequency of the G allele for rs1799971 has been reported to be 12.2%, there are considerable differences between ethnicities, with Asian populations often demonstrating an increased alternate G allele frequency (approximately 40%; TogoVarID: tgv27548008, Additional file 2: Table S1). This was supported by the fact that the G allele frequency for rs1799971 in Japanese samples ranges from 42% to 44.7% (Additional file 2: Table S1). Here, we detected the G allele in two of the five Japanese study participants, which was consistent with previous observations. SNPs rs1045642 [18, 19] and rs2165870 [20, 21] also experience a moderate-to-high alternate allele frequency in the Japanese population, with frequencies of approximately 60% and 70%, respectively. This was consistent with our nanopore-based SNP genotyping which identified an alternate G allele for rs1045642 in three of the five Japanese participants and the rs2165870 alternate G allele in all five samples. In contrast to these findings, public datasets suggest that the alternate allele frequencies for rs4369876 [22], rs33985936 [23], and rs140124801 [24] are quite low. This was also the case in our study where all but one individual, with the alternate A allele for rs4369876, presented as homozygous for the major reference allele at all three SNP loci. Thus, taken together these results suggest that nanopore mediated SNP genotyping accurately reflects the common allele frequencies associated with these SNPs in the Japanese population, even in this very limited sample.

Conventional NGS generates vast quantities of highly accurate sequencing data on a large scale [13, 25]. However, the cost-effectiveness of these platforms are dependent on the acquisition and evaluation of large numbers of samples to be analyzed in one batch. Nanopore sequencing enables the real-time processing of a small number of samples on a case-by-case basis, making this analysis more suitable for genetic testing in personalized clinical care as opposed to large-scale clinical studies [26, 27]. Given these features, we designed this study to establish and evaluate a workflow for rapid SNP genotyping on a per patient basis, and although this study focused on SNPs linked to perioperative outcomes, nanopore amplicon sequencing and a similar bioinformatics pipeline could easily be applied in the detection of a wide variety of genetic variations. Despite the obvious advantages presented by this approach our study did suffer from several limitations. First, the reliability of this method was only evaluated for a very small number of known SNPs in a very small population. Second, this genotyping workflow limits the identification of novel or unknown SNPs. Third, the clinical relevance of each SNP has not yet been thoroughly validated. Accumulating evidence has shown the potential of several SNPs to act as genetic markers for risk prediction around perioperative outcomes. However, the utility of SNP genotyping data in anesthetic management remains unclear. Further studies are required to elucidate the underlying mechanisms linking these genetic variations to the clinical phenotype and to establish a basis for the practice of precision medicine in perioperative care.

## Conclusion

Nanopore sequencing reads were reasonably well clustered within their expected genotypes, and this simplified workflow provided reliable allele frequency estimation with a total analysis time of less than 4 h. This suggests that nanopore sequencing may be a promising and versatile tool for SNP genotyping, allowing for rapid and reliable perioperative risk prediction in clinical settings.

## Data Availability

The data used in this study is available from the corresponding author upon reasonable request.

## Abbreviations

NGS: next-generation sequencing
PCR: polymerase chain reaction
SNP: single nucleotide polymorphism
SNV: single nucleotide variation

## Supplementary Information

**Additional file 1** Bioinformatics analysis.

**Additional file 2: Table S1** Allele frequency data for selected SNPs associated with perioperative outcomes

**Additional file 3: Figs. S1-S6** SNP genotyping using dideoxy sequencing.

**Additional file 4: Table S2** Statistics of the nanopore sequencing data.

**Additional file 5: Tables S3-S8** Allele frequencies for each SNP as determined by nanopore sequencing.

## Declarations

### Ethics approval and consent to participate

This study was approved by the Institutional Review Board at Kansai Medical University Hospital (No. 2020285, February 19, 2021).

### Consent for publication

Not applicable.

### Availability of data and materials

The data used in this study is available from the corresponding author upon reasonable request.

### Authors’ contributions

YM and KH designed and supervised this study. YT and AO assisted with sample collection. YT and YM conducted the experiments. YT, YM, and YF analyzed the data. YM wrote the manuscript. YT and KH contributed to the manuscript editing. All the authors have read and approved the final manuscript.

## Acknowledgements

We would like to thank Editage (www.editage.com) for English language editing.

