## Additional file 1 for "Rapid detection of single nucleotide polymorphisms using the MinION nanopore sequencer: a feasibility study for perioperative precision medicine"

### Bioinformatics analysis

#### The exact commands used to generate data

**Step 1** Extraction of reads with a perfect match to the probe sequence

- Input: seq.fastq (pass reads)
- Output: seq_1.fastq

seqkit grep -s -d -p (probe sequence) seq.fastq > seq_1.fastq

**Step 2** Random sampling (1000 reads)

- Input: seq_1.fastq
- Output: seq_2.fastq

seqkit sample -n 1000 -2 seq_1.fastq > seq_2.fastq

**Step 3** Estimation of allele frequencies

- Input: seq_2.fastq
- Output: read counts

seqkit grep -s -d -p (allele sequence) seq_2.fastq | seqkit seq -n | wc -l

**Sequence information**

**rs1045642**

probe sequence: CCTCACNATCTCT
reference allele: CCTCACAATCTCT
alternate allele: CCTCACGATCTCT
others: CCTCACYATCTCT

**rs1799971**

probe sequence: TAGATGGCNACCT
reference allele: TAGATGGCAACCT
alternate allele: TAGATGGCGACCT
others: TAGATGGCYACCT

**rs2165870**

probe sequence: AGCCTGNTATACT
reference allele: AGCCTGATATACT
alternate allele: AGCCTGGTATACT
others: AGCCTGYTATACT

**rs4369876**

probe sequence: TTTCANATAATTT
reference allele: TTTCACATAATTT
alternate allele: TTTCAAATAATTT
others: TTTCAKATAATTT

**rs33985936**

probe sequence: CATGCCTGANGCC
reference allele: CATGCCTGACGCC
alternate allele: CATGCCTGATGCC
others: CATGCCTGARGCC

**rs140124801**

probe sequence: CGTAGCCCANCGT
reference allele: CGTAGCCCACCGT
alternate allele: CGTAGCCCATCGT
others: CGTAGCCCARCGT

**Example**

For genotyping of SNP rs1045642:

**Step 1** Extraction of reads with a perfect match to the probe sequence

```
seqkit grep -s -d -p CCTCACNATCTCT seq.fastq > seq_1.fastq
```

**Step 2** Random sampling (1000 reads)

```
seqkit sample -n 1000 -2 seq_1.fastq > seq_2.fastq
```

**Step 3** Estimation of allele frequencies

Reference allele (A):

```
seqkit grep -s -d -p CCTCACAATCTCT seq_2.fastq | seqkit seq -n | wc -l
```

Alternate allele (G):

```
seqkit grep -s -d -p CCTCACGATCTCT seq_2.fastq | seqkit seq -n | wc -l
```

Others (C/T):

```
seqkit grep -s -d -p CCTCACYATCTCT seq_2.fastq | seqkit seq -n | wc -l
```
