## Additional file 2 for "Rapid detection of single nucleotide polymorphisms using the MinION nanopore sequencer: a feasibility study for perioperative precision medicine"

**Table S1** Allele frequency data for selected SNPs associated with perioperative outcomes

| SNP | Variant | Alternate allele frequency (%) | | | | | |
|---|---|---|---|---|---|---|---|
| | Ref>Alt | Japanese genetic variation database | | | | gnomAD Genomes | |
| | | GEM-J | JGA-SNP | ToMMo | HGVD | East Asian | Total |
| rs1045642 | A>G | 58.3 | 57.7 | 58.3 | 59.9 | 63.1 | 56.0 |
| rs1799971 | A>G | 44.3 | 44.7 | 44.7 | 42.0 | 33.7 | 12.2 |
| rs2165870 | A>G | 74.1 | NA | 73.7 | NA | 84.6 | 75.6 |
| rs4369876 | C>A | 4.5 | 4.4 | 4.4 | 5.3 | 5.7 | 1.0 |
| rs33985936 | C>T | 8.5 | 8.4 | 8.2 | 9.3 | 12.5 | 20.6 |
| rs140124801 | C>T | NA | NA | NA | NA | 0 | 0.5 |

Ref: reference allele, Alt: alternate allele, NA: not available. Data are from the integrated genome variation database TogoVar[1]. Allele frequency data of Japanese population from the public databases (GEM-J[2], JGA-SNP[3], ToMMo[4], and HGVD[5]) are shown. The data for east Asian and total populations (African-American/African, Ashkenazi Jewish, East Asian, Finnish, Latino, Non-Finnish European, and Other) are obtained from gnomAD[6].


1. TogoVar: Tokyo: Japan Science and Technology Agency (Japan), National Bioscience Database Center. TogoVar ID tgv30006530 (rs1045642), tgv27548008 (rs1799971), tgv5354698 (rs2165870), tgv9362554 (rs4369876), tgv12122442 (rs33985936), tgv56421148 (rs140124801); [2021/12/18]. Available from: https://togovar.biosciencedbc.jp
2. GEM-J: "GEM Japan Whole Genome Aggregation (GEM-J WGA) Panel". Japan: GEnome Medical alliance Japan Project (GEM-J). Available from: https://togovar.biosciencedbc.jp/doc/datasets/gem_j_wga
3. JGA-SNP: Variant set aggregated from SNP-chip data in NBDC Human Database/JGA. Tokyo: Japan Science and Technology Agency (Japan), National Bioscience Database Center. JGA-SNP dataset; [2021/12/18]. Available from: https://togovar.biosciencedbc.jp/doc/datasets/jga_ngs
4. ToMMo: Tohoku Medical Megabank Organization 8.3K JPN Allele Frequency Panel.
   Tadaka et al., "jMorp updates in 2020: large enhancement of multi-omics data resources on the general Japanese population", Nucleic Acids Research. 2021 Jan 8;49(D1):D536-D544. https://doi.org/10.1093/nar/gkaa1034
5. HGVD: The Human Genetic Variation Database.
   K. Higasa et al. "Human genetic variation database, a reference database of genetic variations in the Japanese population" J Hum Genet 61:547-553, 2016. https://doi.org/10.1038/jhg.2016.12
6. gnomAD: The Genome Aggregation Database. Karczewski, K.J., Francioli, L.C., Tiao, G. et al. The mutational constraint spectrum quantified from variation in 141,456 humans. Nature 581, 434–443 (2020). https://doi.org/10.1038/s41586-020-2308-7
