## Additional file 3 for "Rapid detection of single nucleotide polymorphisms using the MinION nanopore sequencer: a feasibility study for perioperative precision medicine"

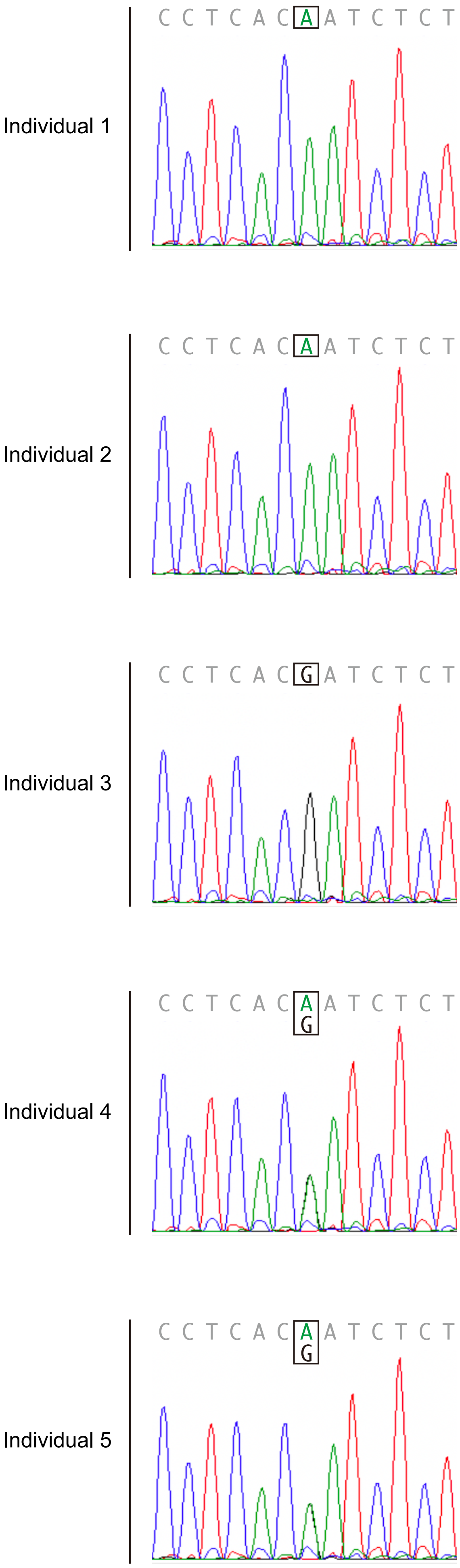


**Fig. S1** Genotyping of SNP rs1045642 by the dideoxy sequencing. Representative electropherograms of five individuals are shown: 1, A/A homozygote; 2, A/A homozygote; 3, G/G homozygote; 4, A/G heterozygote; 5, A/G heterozygote. The bases corresponding to the SNP site are boxed.

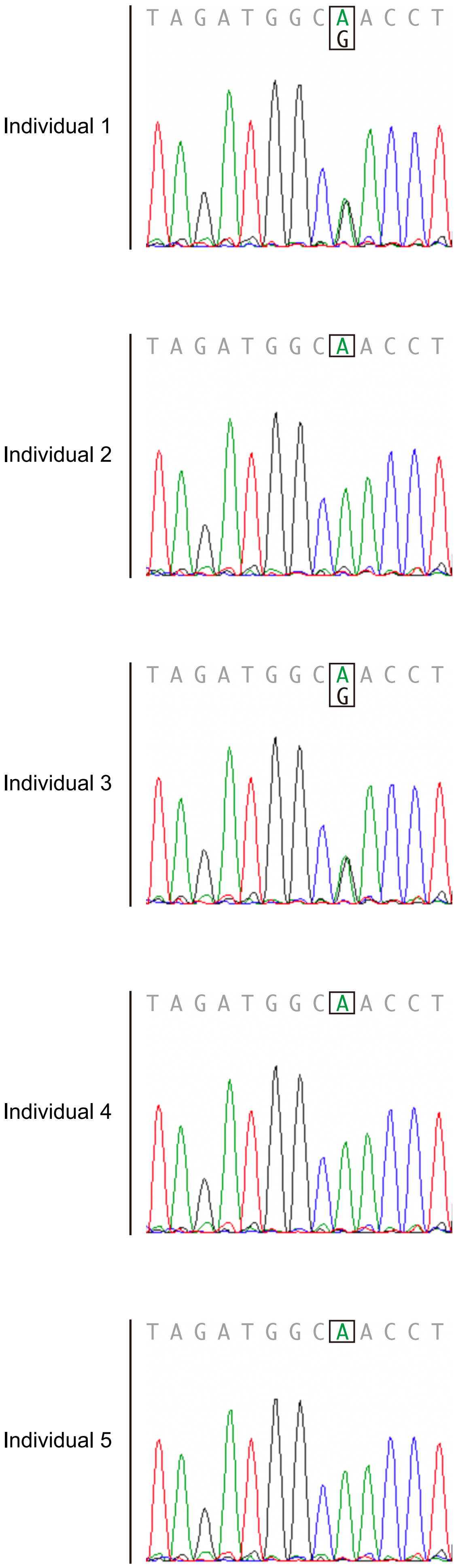


**Fig. S2** Genotyping of SNP rs1799971 by the dideoxy sequencing. Representative electropherograms of five individuls are shown: 1, A/G heterozygote; 2, A/A homozygote; 3, A/G heterozygote; 4, A/A homozygote; 5, AA homozygote. The bases corresponding to the SNP site are boxed.

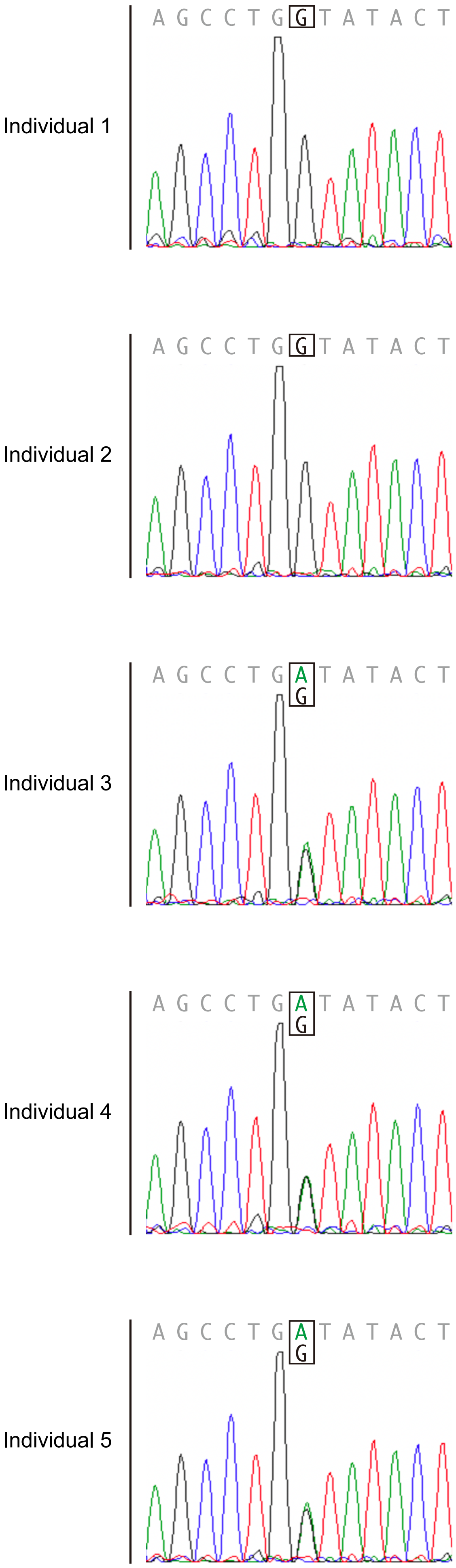


**Fig. S3** Genotyping of SNP rs2165870 by the dideoxy sequencing. Representative electropherograms of five individuals are shown: 1, G/G homozygote; 2, G/G homozygote; 3, A/G heterozygote; 4, A/G heterozygote; 5, A/G heterozygote. The bases corresponding to the SNP site are boxed.

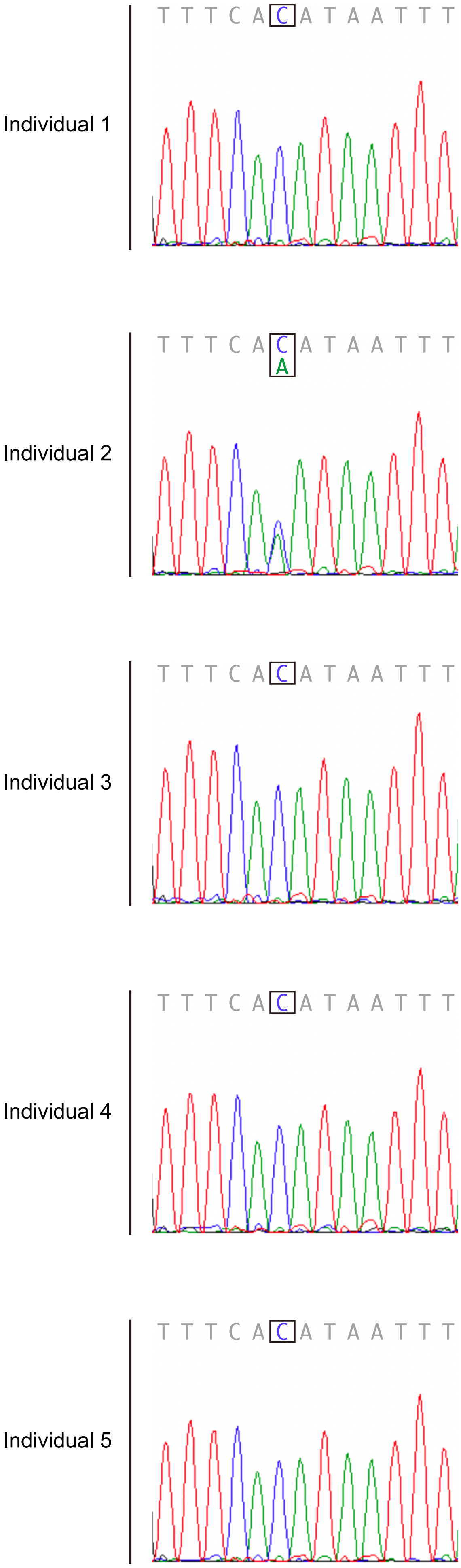


**Fig. S4** Genotyping of SNP rs4369876 by the dideoxy sequencing. Representative electropherograms of five individuals are shown: 1, C/C homozygote; 2, C/A heterozygote; 3, C/C homozygote; 4, C/C homozygote; 5, C/C homozygote. The bases corresponding to the SNP site are boxed.

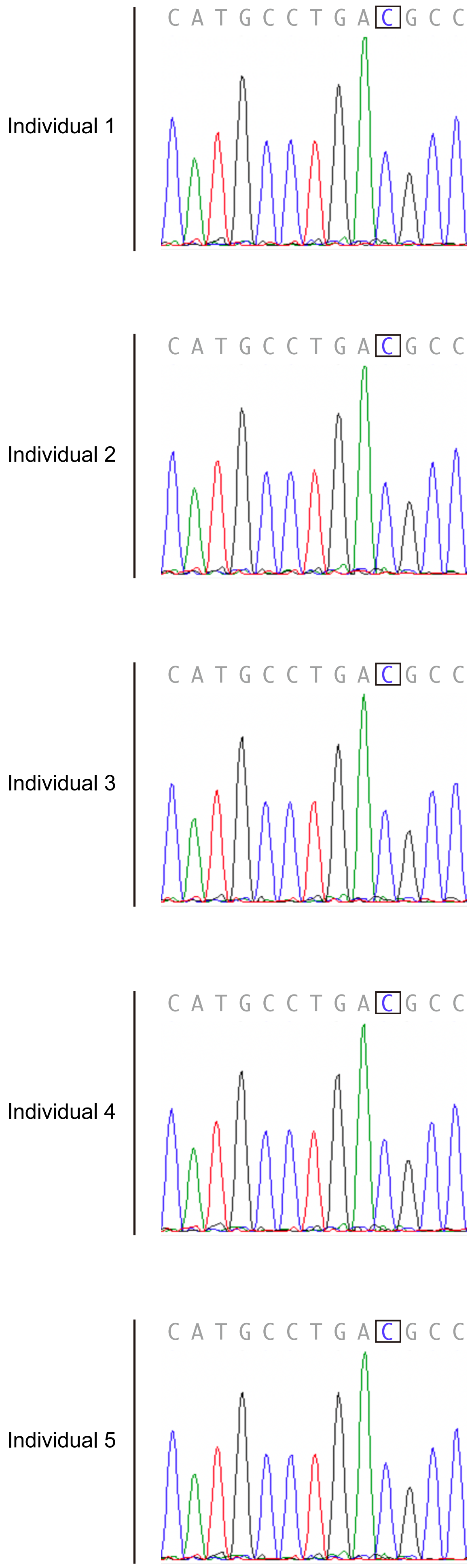


**Fig. S5** Genotyping of SNP rs33985936 by the dideoxy sequencing. Representative electropherograms of five individuals are shown: 1, C/C homozygote; 2, C/C homozygote; 3, C/C homozygote; 4, C/C homozygote; 5, C/C homozygote. The bases corresponding to the SNP site are boxed.

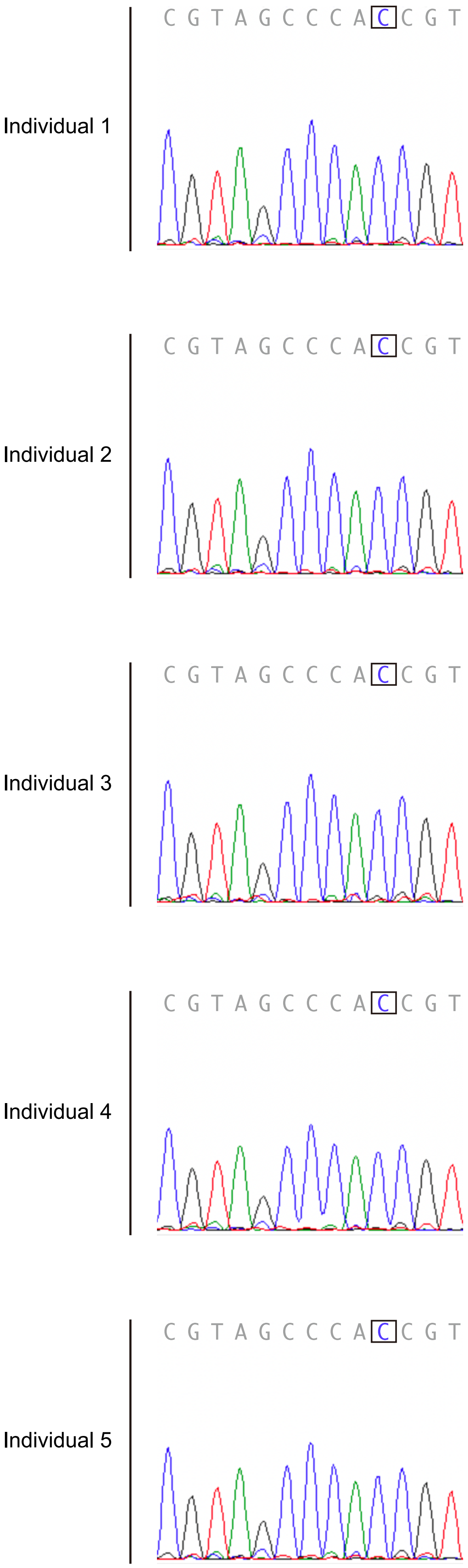


**Fig. S6** Genotyping of SNP rs140124801 by the dideoxy sequencing. Representative electropherograms of five Individuals are shown: 1, C/C homozygote; 2, C/C homozygote; 3, C/C homozygote; 4, C/C homozygote;
