## Additional file 4 for "Rapid detection of single nucleotide polymorphisms using the MinION nanopore sequencer: a feasibility study for perioperative precision medicine"

**Table S2** Statistics of the nanopore sequencing data

| SNP | | Avg (bp) | Q score |
|---|---|---|---|
| rs1045642 | Individual 1 | 414.5 | 12.5 |
| | Individual 2 | 428.1 | 12.5 |
| | Individual 3 | 416.4 | 12.7 |
| | Individual 4 | 418.4 | 11.5 |
| | Individual 5 | 429.0 | 9.4 |
| rs1799971 | Individual 1 | 714.2 | 11.8 |
| | Individual 2 | 733.7 | 12.2 |
| | Individual 3 | 723.3 | 12.0 |
| | Individual 4 | 726.0 | 11.1 |
| | Individual 5 | 723.9 | 9.2 |
| rs2165870 | Individual 1 | 921.0 | 9.5 |
| | Individual 2 | 929.3 | 13.2 |
| | Individual 3 | 891.6 | 13.0 |
| | Individual 4 | 906.0 | 12.0 |
| | Individual 5 | 929.1 | 9.5 |
| rs4369876 | Individual 1 | 432.2 | 14.8 |
| | Individual 2 | 444.8 | 13.5 |
| | Individual 3 | 434.6 | 13.3 |
| | Individual 4 | 436.7 | 12.1 |
| | Individual 5 | 440.0 | 11.7 |
| rs33985936 | Individual 1 | 463.7 | 13.8 |
| | Individual 2 | 471.9 | 12.4 |
| | Individual 3 | 464.3 | 13.9 |
| | Individual 4 | 460.4 | 13.8 |
| | Individual 5 | 467.5 | 10.9 |
| rs140124801 | Individual 1 | 482.4 | 13.3 |
| | Individual 2 | 480.3 | 13.3 |
| | Individual 3 | 477.0 | 13.3 |
| | Individual 4 | 481.8 | 13.4 |
| | Individual 5 | 474.0 | 12.0 |

Statistics of 1000 nanopore sequencing reads used for SNP genotyping.

Avg: average read length, Q score: average Phred quality score.
