## Additional file 5 for "Rapid detection of single nucleotide polymorphisms using the MinION nanopore sequencer: a feasibility study for perioperative precision medicine"

**Table S3** Allele frequencies for rs1045642 determined by nanopore sequencing

| | Read count | | | | Allele frequency (%) | | |
|---|---|---|---|---|---|---|---|
| Genotype | A | G | C/T | Total | A | G | C/T |
| Individual 1 | 994 | 5 | 1 | 1000 | 99.4 | 0.5 | 0.1 |
| Individual 2 | 998 | 1 | 1 | 1000 | 99.8 | 0.1 | 0.1 |
| Individual 3 | 1 | 998 | 1 | 1000 | 0.1 | 99.8 | 0.1 |
| Individual 4 | 464 | 530 | 6 | 1000 | 46.4 | 53.0 | 0.6 |
| Individual 5 | 463 | 537 | 0 | 1000 | 46.3 | 53.7 | 0 |

**Table S4** Allele frequencies for rs1799971 determined by nanopore sequencing

| | Read count | | | | Allele frequency (%) | | |
|---|---|---|---|---|---|---|---|
| Genotype | A | G | C/T | Total | A | G | C/T |
| Individual 1 | 446 | 554 | 0 | 1000 | 44.6 | 55.4 | 0 |
| Individual 2 | 993 | 7 | 0 | 1000 | 99.3 | 0.7 | 0 |
| Individual 3 | 435 | 565 | 0 | 1000 | 43.5 | 56.5 | 0 |
| Individual 4 | 989 | 8 | 3 | 1000 | 98.9 | 0.8 | 0.3 |
| Individual 5 | 994 | 6 | 0 | 1000 | 99.4 | 0.6 | 0 |

**Table S5** Allele frequencies for rs2165870 determined by nanopore sequencing

| | Read count | | | | Allele frequency (%) | | |
|---|---|---|---|---|---|---|---|
| Genotype | A | G | C/T | Total | A | G | C/T |
| Individual 1 | 5 | 985 | 10 | 1000 | 0.5 | 98.5 | 1.0 |
| Individual 2 | 5 | 990 | 5 | 1000 | 0.5 | 99.0 | 0.5 |
| Individual 3 | 488 | 507 | 5 | 1000 | 48.8 | 50.7 | 0.5 |
| Individual 4 | 498 | 498 | 4 | 1000 | 49.8 | 49.8 | 0.4 |
| Individual 5 | 580 | 416 | 4 | 1000 | 58.0 | 41.6 | 0.4 |

**Table S6** Allele frequencies for rs4369876 determined by nanopore sequencing

| | Read count | | | | Allele frequency (%) | | |
|---|---|---|---|---|---|---|---|
| Genotype | C | A | G/T | Total | C | A | G/T |
| Individual 1 | 981 | 0 | 19 | 1000 | 98.1 | 0 | 1.9 |
| Individual 2 | 461 | 530 | 9 | 1000 | 46.1 | 53.0 | 0.9 |
| Individual 3 | 979 | 0 | 21 | 1000 | 97.9 | 0 | 2.1 |
| Individual 4 | 971 | 0 | 29 | 1000 | 97.1 | 0 | 2.9 |
| Individual 5 | 943 | 0 | 57 | 1000 | 94.3 | 0 | 5.7 |

**Table S7** Allele frequencies for rs33985936 determined by nanopore sequencing

| | Read count | | | | Allele frequency (%) | | |
|---|---|---|---|---|---|---|---|
| Genotype | C | T | A/G | Total | C | T | A/G |
| Individual 1 | 992 | 8 | 0 | 1000 | 99.2 | 0.8 | 0.0 |
| Individual 2 | 990 | 10 | 0 | 1000 | 99.0 | 1 | 0.0 |
| Individual 3 | 994 | 6 | 0 | 1000 | 99.4 | 0.6 | 0.0 |
| Individual 4 | 992 | 8 | 0 | 1000 | 99.2 | 0.8 | 0.0 |
| Individual 5 | 993 | 7 | 0 | 1000 | 99.3 | 0.7 | 0.0 |

**Table S8** Allele frequencies for rs140124801 determined by nanopore sequencing

| | Read count | | | | Allele frequency (%) | | |
|---|---|---|---|---|---|---|---|
| Genotype | C | T | A/G | Total | C | T | A/G |
| Individual 1 | 998 | 1 | 1 | 1000 | 99.8 | 0.1 | 0.1 |
| Individual 2 | 1000 | 0 | 0 | 1000 | 100 | 0 | 0 |
| Individual 3 | 996 | 4 | 0 | 1000 | 99.6 | 0.4 | 0 |
| Individual 4 | 999 | 1 | 0 | 1000 | 99.9 | 0.1 | 0 |
| Individual 5 | 998 | 1 | 1 | 1000 | 99.8 | 0.1 | 0.1 |
